# Serial evaluation of anti-SARS-CoV-2 IgG antibody and breakthrough infections in BNT162b2 Vaccinated migrant workers from Bangladesh

**DOI:** 10.1101/2021.09.07.21263221

**Authors:** Ashraful Hoque, Md Marufur Rahman, Anindita Das

**Affiliations:** Department of Blood Transfusion, Sheikh Hasina National Institute of Burn and Plastic Surgery, Dhaka; Centre for Medical Biotechnology, Management Information System, Directorate General of Health Services, Dhaka; Department of Internal Medicine, Dhaka Medical College Hospital, Dhaka

## Abstract

**Background:** While Bangladesh has started its mass COVID-19 vaccination drive, it is struggling to cover its huge population similar to other low- and middle-income countries due to the lack of vaccine availability. One of the major remittance sources for Bangladesh is its migrant workers who are required to receive mRNA vaccines to return to their jobs. Despite reports of higher efficacy of mRNA vaccine against COVID-19, breakthrough infection cases are arising especially with the emergence of Delta variant. It is highly important to understand the post-vaccination immune response and breakthrough infections in different populations so that the necessity of booster dosage can be assessed properly.

**Methods:** We observed post BNT162b2 full vaccination immune response in a small older group (mean age= 59.5±5.44 years) of migrant workers (n=10) for six months at the Sheikh Hasina National Institute of Burn and Plastic Surgery, Dhaka, Bangladesh. The plasma samples from the participants were collected after 14 days, 2 months, 3 months, 4 months, 5 months, and 6 months of receiving the 2^nd^ dose of the BNT162b2 vaccine. Anti S1 RBD IgG responses were measured as optical density ratios using a commercially available ELISA kit.

**Results:** All 10 of the participants were male migrant workers and none of them had a history of previous COVID-19 infection. The median antibody response [IQ1:IQ3] was 9.05 [7.53; 10.0] on day 14 then it increases to 13.6 [12.0; 14.0] at the second month which gradually decreased to a median of 8.63 [8.34; 9.37] on the 6th-month post-vaccination. There were two breakthrough infection cases after receiving the second dose and the antibody responses were highly increased in the following months. Two of the breakthrough cases were diagnosed with mild COVID-19 as the symptom duration was less than 3 days with no respiratory complications and no hospital admission were required.

**Conclusions:** The BNT162b2 mRNA vaccine produces a strong immune response that sustains at least 6 months after getting fully vaccinated. But even after getting fully vaccinated people are susceptible to breakthrough infections that are not severe and boost the immune response greatly offering a hybrid immunity from both vaccine and natural infection. Hence, it is still important to fully vaccinate a greater number of people rather than thinking of offering booster dosage to a privileged population out of the fear of breakthrough cases. If the LMICs can quickly cover at least 80% of their population with usual priority targets (healthcare workers, migrant workers, older people, etc.) then a global risk reduction and pandemic control would be possible that will allow additional variant-specific boosters for targeted populations if evidence support.

## Introduction

Countries around the world are now racing to vaccinate people against SARS-CoV-2, the virus that causes COVID-19. It is one of the most ambitious vaccination programs ever. To bring this epidemic to an end, a large population must be immune to the virus. Vaccination is one of the most secure approaches to accomplish this. Vaccines are a technique on which humankind has frequently relied in the past to reduce the mortality toll from infectious diseases.

Rapid vaccine-induced population immunity is a critical global strategy for COVID-19 pandemic control. Vaccination efforts must maximize their early effects, especially given the rapid spread of novel variants. According to the World Health Organization vaccination dashboard, a total of 3,946,458,313 vaccination doses has been administered as of August 4, 2021 (1). Bangladesh began administering COVID-19 vaccination on January 27, 2021, while mass vaccination began on February 7, 2021. So far, the number of the first dose administered is 10,009,953 and for the second the number is 4,416,131 till August 5, 2021 (2). Bangladesh has a huge number of working population living outside the country especially in the middle east. They are the largest source of remittance for the country and have a huge impact on the country’s economy. A lot of them returned to the country during this year and had received Pfizer– BioNTech COVID-19 mRNA vaccine (BNT162b2) known to have excellent efficacy against the virus compared to other COVID-19 vaccines (3,4)

Most of the SARS-CoV-2 vaccines have been developed to induce the production of antibodies against the SARS-CoV-2 spike protein. Admittedly, effective COVID-19 vaccination requires more than merely finishing the vaccine doses. The efficacy of the COVID-19 vaccine can significantly depend on sufficient production of SARS COV-2 neutralizing antibodies and persistence of the antibodies over time. Therefore, the serial measurement of circulating anti-S-RBD IgG levels may provide precious information on acquired immunity against SARS-CoV-2. The frequency of doses and requirement of booster doses may also be determined from such experiments. However, several studies that were primarily performed on booster responses to vaccines have reported that the immune responses were less effective in some people especially at old age when immune-senescence develops. Lower antibody concentrations, with higher decline rates below seroprotective margins in such a population, may result in an overall shortened period of protection (5–7). Hence it is important to know how long the antibodies persist after vaccination in a specific population and how the efficacy changes especially with the emergence of newer variants as a manifestation of breakthrough cases.

We examined SARS-CoV-2-specific antibody responses in a small group of migrant workers - from Bangladesh after completion of both doses of the BNT162b2 vaccine to identify the duration of antibody response over a time period (six months) and occurrences of breakthrough cases.

## Materials and methods

This observational study was conducted at Sheikh Hasina National Institute of Burn and Plastic Surgery, Dhaka between January 10, 2021, to August 07, 2021. We enrolled 10 participants for our study who received both doses of the BNT162b2 vaccine. Participants’ demographic data along with past SARS-COV-2 infection history and existing comorbidities were recorded using a structured questionnaire. The plasma samples from the participants were collected after 14 days, 2 months, 3 months, 4 months, 5 months, and 6 months of receiving the 2^nd^ dose. Serological testing for antibodies to the RBD of the S1 subunit of the viral spike protein (IgG) was performed from the plasma samples using a commercially available Anti-SARS-CoV-2 ELISA kit as per instruction supplied by the manufacturer. The study protocol was reviewed and approved by the Institutional Review Committee of the Sheikh Hasina National Institute of Burn and Plastic Surgery, Dhaka (approval memo number: SHNIBPS/Jan-14(09)). Data were summarized by descriptive analysis. Numerical variables are reported as mean and standard deviation or median and interquartile ranges, as appropriate. Categorical variables are reported as counts and percentages. The analysis was performed in Graphpad Prism 9.0.

## Results

### Demographics and clinical characteristics of the patients

Plasma samples were collected from 10 individuals who received two complete doses of BNT162b2 mRNA Covid-19 Vaccine. The demographic and clinical characteristics of the participants are shown in Table 1. The mean (±SD) age of participants was 59.5±5.44 years (range, 48 to 67); 100% were men. None of the participants were previously infected with the SARS-CoV-2 virus. Almost third of patients (4 [40%]) had no comorbidities while the other two thirds (6 [60%]) had hypertension; (3 [30%]) had lone hypertension, (1 [10%]) had hypertension and biliary atresia, (1 [10%]) had hypertension and chronic obstructive lung disease (COPD), and (1 [10%]) have hypertension and diabetes mellitus.

**Table 1.** Demographics and clinical characteristics of participants.

| Characteristics | Patients (N=10) |
| --- | --- |
| Age | 59.5 (±5.44) years |
| Sex |  |
| Male | 10 (100%) |
| Previously infected |  |
| YES | 0 (0%) |
| No | 10 (100%) |
| Comorbidity |  |
| Absent | 4 (40.0%) |
| Hypertension | 6 (60.0%) |
| Bronchial Asthma | 1 (10.0%) |
| COPD | 1 (10.0%) |
| Diabetes mellitus | 1 (10.0%) |

### BNT162b2 mRNA Covid-19 Vaccine antibody response

Plasma samples were collected at 6 time periods after the Covid-19 vaccine 2^nd^ dose; 14 days, 2 months, 3 months, 4 months, 5 months, and 6 months. The antibody response was measured and compared as Optical Density Ratio (ODR) from ELISA readings. The median and interquartile ranges of antibody levels (ODR) over 6 month periods are shown in Table 2, and Figure 1. The antibody response median [IQ1:IQ3] is 9.05 [7.53; 10.0] at day 14 then it increases to 13.6 [12.0; 14.0] at the second month. However, it shows a gradual decrease over the third- and fourth-month periods with medians [IQ1:IQ3] 11.9 [11.2; 12.6] and 10.3 [10.0; 11.0]. The antibody response median is steady over the fifth and sixth months with levels 9.80 [9.06; 10.1] and 8.63 [8.34; 9.37]. The individual antibody response fluctuations over 6 months are shown in Figure 2. It shows an initial increase of antibody response over the first 2 months and a gradual decrease over the next 4 months in all cases except cases 1 and 2 who showed a remarkable increase from the 4^th^ month to the 5^th^ month.

**Table 2.** BNT162B2 mRNA Covid-19 vaccine antibody titer over 6 month’s summary table.

| **Time** | **Antibody Titer (median [IQ1:IQ3])** |
| --- | --- |
| 14 days | 9.05 [7.53;10.0] |
| 2 months | 13.6 [12.0;14.0] |
| 3 months | 11.9 [11.2;12.6] |
| 4 months | 10.3 [10.0;11.0] |
| 5 months | 9.80 [9.06;10.1] |
| 6 months | 8.63 [8.34;9.37] |

| Time | Antibody Titer (median [IQ1;IQ3]) |
| --- | --- |
| 14 days | 9.05 [7.53;10.0] |
| 2 months | 13.6 [12.0;14.0] |
| 3 months | 11.9 [11.2;12.6] |
| 4 months | 10.3 [10.0;11.0] |
| 5 months | 9.80 [9.06;10.1] |
| 6 months | 8.63 [8.34;9.37] |

**Figure 1.**
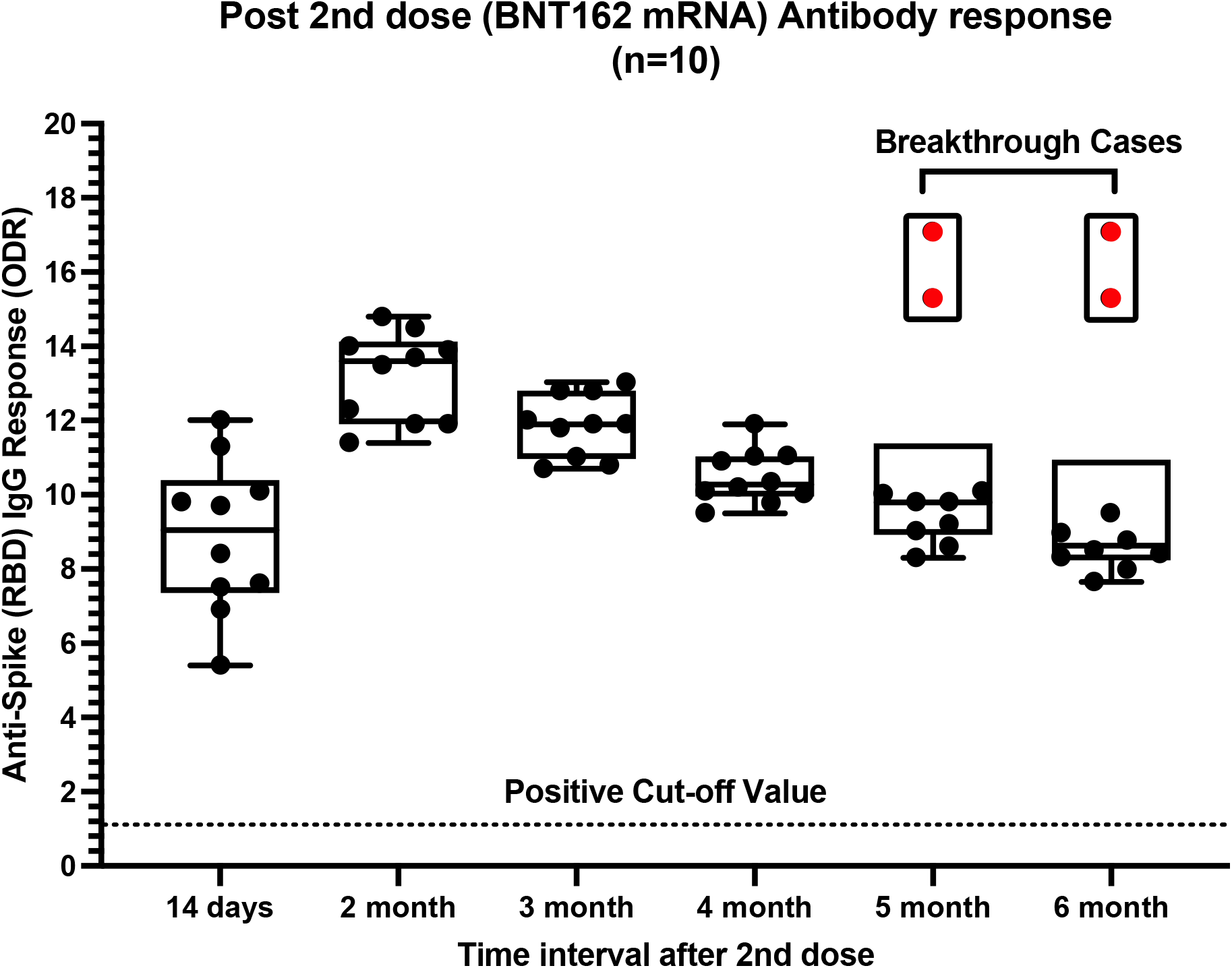
Post BTN162b2 (2^nd^ dose) antibody response over 6 months. Boxplot for BNT162B2 mRNA Covid-19 vaccine antibody response (ODR) median and interquartile ranges over 6 months

**Figure 2.**
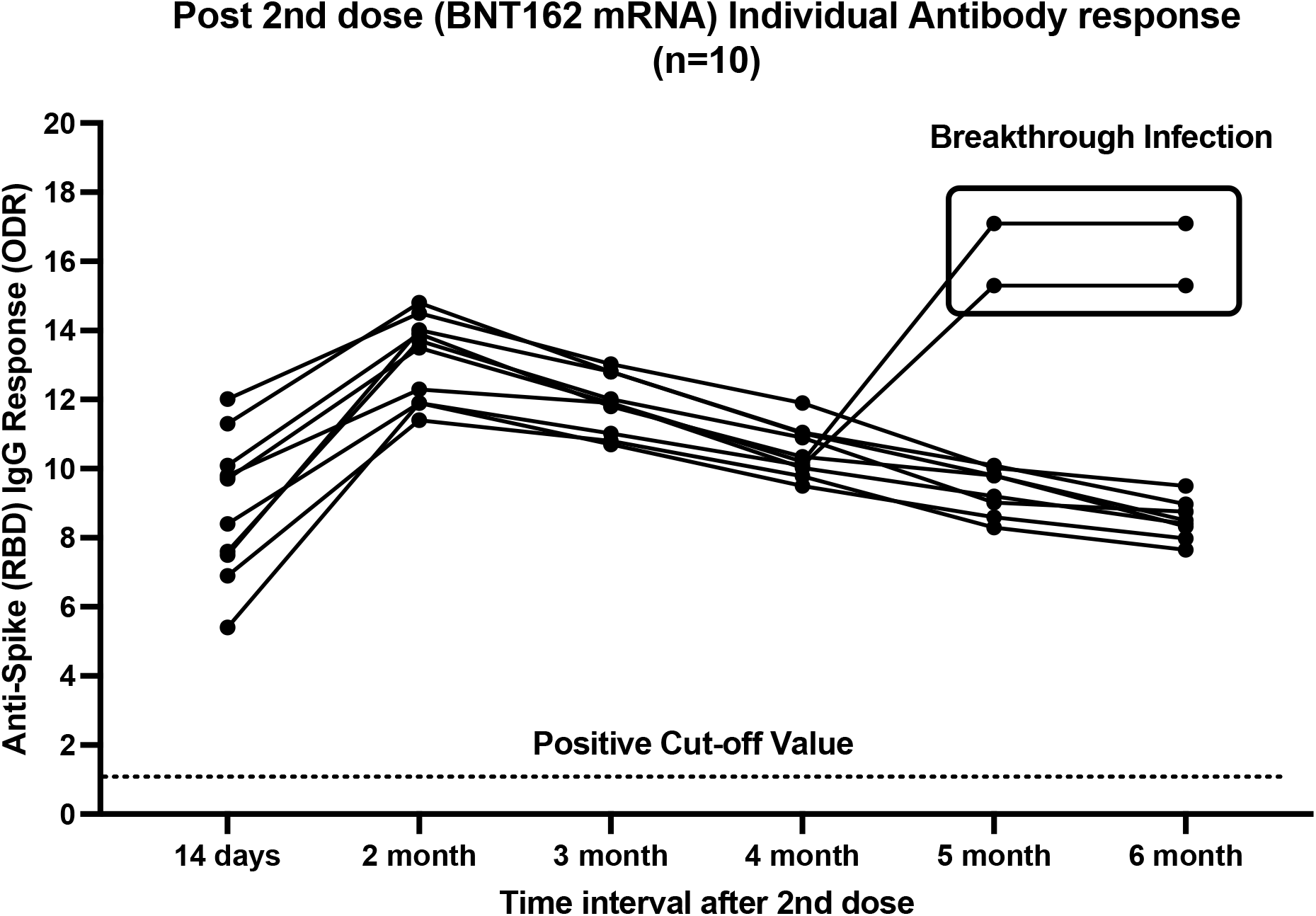
Post BTN162b2 (2^nd^ dose) individual antibody response over 6 months. BNT162B2 mRNA Covid-19 vaccine antibody response (ODR) over 6 months after the 2^nd^ dose by individual cases

### Covid-19 Breakthrough cases

SARS-CoV-2 RNA was detected in the respiratory specimen of 2 cases among 10 participants after 84- and 91-days post-vaccination. The two cases are males aged 61 and 63 years old. Both of them had a fever for less than three days and the disease was categorized as mild as respiratory symptoms were absent with SpO2 within the normal limit. No hospital admission was required. Their antibody response trend line was significantly different from other cases over months 4 and 5 with a remarkable increase compared to other cases in which their lines were continuing to decrease gradually as shown in Figure 2.

## Discussion & Conclusion

As the majority of our migrant workers from Bangladesh are men all of our study participants were in the same gender group (8). Our observation revealed successful antibody response after receiving two doses of BNT162 mRNA vaccine which increased till 2^nd^ month then it gradually depleted and reached near baseline at 6^th^ month. Though the anti-RBD S1 IgG level was declining it was still found at six-month post-vaccination in all of our participants. Similar findings were observed in other studies for both BNT162b2 and another mRNA-based COVID-19 vaccine mRNA-1273 (9,10).

While several other studies reported that breakthrough cases are possible after full dose COVID-19 vaccination, the disease remains mild in most cases, we observed the same for the two of our breakthrough cases (11–13). We did not sequence the RNA sample from them but there is a high probability that the cases were infected with the delta variant since Bangladesh was reported to have a delta wave since June with more than 90% of sequenced cases found to be the delta variant (14). Since delta reduces the regular efficacy of COVID vaccines, we might get a high number of breakthrough cases even after full vaccination (4).

Although the direct correlation of vaccine efficacy with antibody titre is still not established, it guides us about the strength of the humoral arm of the immune protection against the virus. Our observation shows antibody response persists at least six months after receiving two doses of BNT162b2. The breakthrough infections boost antibody response further which may be found for an even longer period. A hybrid immunity which is a mix of immune protection from natural infection and vaccination has been discussed earlier by other scientists. Individuals with prior infection generate a stronger immune response after vaccination compared to an immune response from natural infection or vaccination alone (15). Such hybrid immunity can also be observed in breakthrough cases which we already have found in our observations. Studies identified that the antibodies generated from natural infection undergo affinity maturations where they accumulate somatic mutations and provide a larger breadth of protection against multiple variants of concerns (16,17).

Then there is also cellular immunity and memory B-cells which last longer than the antibodies and have wider target areas on the virus. Studies have shown cellular components of immunity either from natural infection or vaccination persist for a long time and can provide protection against the virus (18,19).

One of the key limitations of our study is the small sample size which does not represent the larger population. That is our findings need to be carefully evaluated and tested further through more extensive research with larger samples. Overall, our observation and findings from other studies suggest COVID-19 vaccines are likely to provide long-term protection. Despite occurrences of breakthrough infections, it’s not worrisome yet as most breakthrough cases are mild even with the current aggressive delta variant prevalence all over the world. The World Health Organization has strongly protested against the decision of giving booster doses as there is a lack of evidence in support of such doses (20). The rise of breakthrough cases is worrying thus we can conduct more studies on them and try to identify who is getting more affected so that they can be prioritized for variant-specific booster dosages if strong evidence arises. As Saha et. al. mentioned the world is seeing vaccine apartheid where richer countries are experimenting with booster shots and the low-middle income countries in the Global South including Bangladesh are struggling to control the case and death numbers from COVID as they couldn’t provide a single shot of COVID vaccine to most of its people (21). We must not advise for booster shots unless stronger evidence arises and an equitable vaccine distribution is ensured throughout the world.

## Data Availability

The raw data for the study are archived at the institutional facility and available on request.

## References

1. WHO (COVID-19) Dashboard With Vaccination Data [Internet]. [cited 2021 Aug 5]. Available from: https://covid19.who.int/?adgroupsurvey=%7Badgroupsurvey%7D&gclid=Cj0KCQjwhr2FBhDbARIsACjwLo24RTJvqwkTryA_XufQoozb9P5RgBatz6VG85AzosLGHBxK_2j4cIkaAu5TEALw_wcB

2. Directorate General of Health Services [Internet]. [cited 2021 Aug 5]. Available from: https://dghs.gov.bd/index.php/bd/component/content/article?layout=edit&id=5649

3. McDonald I, Murray SM, Reynolds CJ, Altmann DM, Boyton RJ. Comparative systematic review and meta-analysis of reactogenicity, immunogenicity, and efficacy of vaccines against SARS-CoV-2. npj Vaccines 2021 61 [Internet]. 2021 May 13 [cited 2021 Aug 15];6(1):1–14. Available from: https://www.nature.com/articles/s41541-021-00336-1

4. Bernal JL, Andrews N, Gower C, Gallagher E, Simmons R, Thelwall S, et al. Effectiveness of Covid-19 Vaccines against the B.1.617.2 (Delta) Variant. https://doi.org/101056/NEJMoa2108891 [Internet]. 2021 Jul 21 [cited 2021 Aug 15];385(7):585–94. Available from: https://www.nejm.org/doi/full/10.1056/NEJMoa2108891

5. Pera A, Campos C, López N, Hassouneh F, Alonso C, Tarazona R, et al. Immunosenescence: Implications for response to infection and vaccination in older people. Maturitas [Internet]. 2015 Sep 1 [cited 2021 Aug 15];82(1):50–5. Available from: http://www.maturitas.org/article/S0378512215006751/fulltext

6. Prelog M. Differential Approaches for Vaccination from Childhood to Old Age. Gerontology [Internet]. 2013 [citd 2021 Aug 15];59(3):230–9. Available from: https://www.karger.com/Article/FullText/343475

7. CA S, R A. B-cell responses to vaccination at the extremes of age. Nat Rev Immunol [Internet]. 2009 [citd 2021 Aug 15];9(3):185–94. Available from: https://pubmed.ncbi.nlm.nih.gov/19240757/

8. Labor force participation rate, female (% of female population ages 15+) (modeled ILO estimate) - Bangladesh | Data [Internet]. [cited 2021 Aug 15]. Available from: https://data.worldbank.org/indicator/SL.TLF.CACT.FE.ZS?locations=BD

9. Doria-Rose N, Suthar MS, Makowski M, O’Connell S, McDermott AB, Flach B, et al. Antibody Persistence through 6 Months after the Second Dose of mRNA-1273 Vaccine for Covid-19. https://doi.org/101056/NEJMc2103916 [Internet]. 2021 Apr 6 [cited 2021 Aug 20];384(23):2259–61. Available from:https://www.nejm.org/doi/full/10.1056/nejmc2103916

10. Ciabattini A, Pastore G, Fiorino F, Polvere J, Lucchesi S, Pettini E, et al. Evidence of SARS-Cov-2-specific memory B cells six months after vaccination with BNT162b2 mRNA vaccine. medRxiv [Internet]. 2021 Jul 14 [cited 2021 Aug 20];2021.07.12.21259864. Available from: https://www.medrxiv.org/content/10.1101/2021.07.12.21259864v1

11. Richard T. Ellison III M. Characterizing COVID-19 Breakthrough Infections. NEJM J Watch [Internet]. 2021 Aug 12 [cited 2021 Aug 20];2021. Available from: https://www.jwatch.org/NA53906/2021/08/12/characterizing-covid-19-breakthrough-infections

12. Hacisuleyman E, Hale C, Saito Y, Blachere NE, Bergh M, Conlon EG, et al. Vaccine Breakthrough Infections with SARS-CoV-2 Variants. https://doi.org/101056/NEJMoa2105000 [Internet]. 2021 Apr 21 [cited 2021 Aug 20];384(23):2212–8. Available from: https://www.nejm.org/doi/full/10.1056/NEJMoa2105000

13. Bergwerk M, Gonen T, Lustig Y, Amit S, Lipsitch M, Cohen C, et al. Covid-19 Breakthrough Infections in Vaccinated Health Care Workers. https://doi.org/101056/NEJMoa2109072 [Internet]. 2021 Jul 28 [cited 2021 Aug 20]; Available from: https://www.nejm.org/doi/full/10.1056/NEJMoa2109072

14. Mallapaty S. Delta threatens rural regions that dodged earlier COVID waves. Nature [Internet]. 2021 Aug 19 [cited 2021 Aug 20];596(7872):325–6. Available from: https://www.nature.com/articles/d41586-021-02146-w

15. Crotty S. Hybrid immunity. Vol. 372, Science. American Association for the Advancement of Science; 2021. p. 1392–3.

16. Wang Z, Muecksch F, Schaefer-Babajew D, Finkin S, Viant C, Gaebler C, et al. Naturally enhanced neutralizing breadth against SARS-CoV-2 one year after infection. Nat 2021 5957867 [Internet]. 2021 Jun 14 [cited 2021 Aug 20];595(7867):426–31. Available from: https://www.nature.com/articles/s41586-021-03696-9

17. Muecksch F, Weisblum Y, Barnes CO, Schmidt F, Schaefer-Babajew D, Wang Z, et al. Affinity maturation of SARS-CoV-2 neutralizing antibodies confers potency, breadth, and resilience to viral escape mutations. Immunity [Internet]. 2021 Aug 10 [cited 2021 Aug 20];54(8):1853-1868.e7. Available from: http://www.cell.com/article/S1074761321002946/fulltext

18. Dan JM, Mateus J, Kato Y, Hastie KM, Yu ED, Facility CE, et al. Immunological memory to SARS-CoV-2 assessed for up to 8 months after infection. Science (80-). 2021 Feb 5;371(6529).

19. Turner JS, O’Halloran JA, Kalaidina E, Kim W, Schmitz AJ, Zhou JQ, et al. SARS-CoV-2 mRNA vaccines induce persistent human germinal centre responses. Nat 2021 5967870 [Internet]. 2021 Jun 28 [cited 2021 Aug 20];596(7870):109–13. Available from: https://www.nature.com/articles/s41586-021-03738-2

20. WHO Director-General’s opening remarks at the media briefing on COVID-19 - 4 August 2021 [Internet]. [cited 2021 Aug 20]. Available from: https://www.who.int/director-general/speeches/detail/who-director-general-s-opening-remarks-at-the-media-briefing-on-covid-4-august-2021

21. Saha S, Tanmoy AM, Tanni AA, Goswami S, Sium SM Al, Saha S, et al. New waves, new variants, old inequity: a continuing COVID-19 crisis. BMJ Glob Heal [Internet]. 2021 Aug 1 [cited 2021 Aug 20];6(8):e007031. Available from: https://gh.bmj.com/content/6/8/e007031

